# Mediterranean diet related metabolite profiles and cognitive performance in a Greek population

**DOI:** 10.1101/2022.09.29.22280504

**Authors:** Christopher Papandreou, Christos Papagiannopoulos, Myrto Koutsonida, Afroditi Kanellopoulou, Georgios Markozannes, Georgios Polychronidis, Andreas G Tzakos, Georgios A. Fragkiadakis, Evangelos Evangelou, Evangelia Ntzani, Ioanna Tzoulaki, Eleni Aretouli, Konstantinos K. Tsilidis

## Abstract

**Background:** Evidence suggests that adherence to the Mediterranean diet (MedDiet) affects human metabolism and may contribute to better cognitive performance. However, the underlying mechanisms are not clear.

**Objective:** We generated a metabolite profile for adherence to MedDiet and evaluated its cross-sectional association with aspects of cognitive performance.

**Methods:** A total of 1,250 healthy Greek middle-aged adults from the Epirus Health Study cohort were included in the analysis. Adherence to the MedDiet was assessed using the 14-point Mediterranean Diet Adherence Screener (MEDAS); cognition was measured using the Trail Making Test, the Verbal Fluency test and the Logical Memory test. A targeted metabolite profiling (n = 250 metabolites) approach was applied, using a high-throughput nuclear magnetic resonance platform. We used elastic net regularized regressions, with a 10-fold cross-validation procedure, to identify a metabolite profile for MEDAS. We evaluated the associations of the identified metabolite profile and MEDAS with cognitive tests, using multivariable linear regression models.

**Results:** We identified a metabolite profile composed of 42 metabolites, mainly lipoprotein subclasses and fatty acids, significantly correlated with MedDiet adherence (Pearson r = 0.35, P-value = 5.5 × 10^−37^). After adjusting for known risk factors and accounting for multiple testing, the metabolite profile and MEDAS were not associated with the cognitive tests.

**Conclusions:** A plasma metabolite profile related to better adherence to the MedDiet was not associated with the tested aspects of cognitive performance, in a middle-aged Mediterranean population.

## Introduction

The Mediterranean diet (MedDiet), characterized by high consumption of olive oil, vegetables, fruits, cereals, nuts, legumes, moderate intake of fish and wine, and low intake of red/processed meats, saturated fat, and sugar [1], may reduce oxidative stress, inflammation and vascular risk factors and thus delay age-related cognitive decline [2, 3]. However, biological mechanisms underlying the complex relationship between MedDiet and cognition still remain elusive [4].

Lately, the MedDiet has been suggested to play an important role in cognitive health and has attracted considerable attention across brain research. A recent systematic review highlighted the necessity for adequate nutritional strategies to prevent cognitive impairment [5], but the results in the current literature are inconsistent. Systematic reviews of observational and intervention studies suggest beneficial effects of the MedDiet on cognitive performance [5-8], while others were inconclusive [9, 10]. In a previous analysis from the Epirus Health Study (EHS), no associations were observed between MedDiet and cognitive function in mostly middle-aged participants [11]. The integration of metabolomics into nutritional studies may not only assist the identification of metabolites that could serve as candidate nutritional biomarkers but also improve the understanding of the biological mechanisms through which diet impacts cognition. Furthermore, identifying metabolite profiles of adherence to dietary patterns may be more clinically meaningful compared to self-reported measures of dietary intake that are prone to measurement error [12].

Several studies have examined the relationship between dietary patterns and circulating metabolite profiles using either mass spectrometry (MS) [13-15] or nuclear magnetic resonance (NMR) [16, 17]. With regards to metabolite profiling of the MedDiet, a previous small study conducted in Northern European patients with coronary heart disease measured 59 metabolites using ^1^H-NMR and found 5 metabolites discriminating between “low” and “high” adherents to the MedDiet [18]. A larger study from Spain using Liquid Chromatography-tandem Mass Spectrometry (LC-MS) identified a metabolic profile of adherence to the MedDiet that was associated with a reduced risk of cardiovascular disease in an older, high-risk population [19]. More recently, 6 plasma metabolites measured by LC-MS were consistently associated with lower global cognitive function and also with adherence to the MedDiet across multi-ethnic studies [20]. However, to the best of our knowledge, no study has assessed the relationship between metabolite profiles of the MedDiet and cognitive performance.

We therefore aimed to analyze plasma metabolites using a high-throughput NMR platform and identify a metabolite profile of adherence to the MedDiet among Greek Middle-aged adults from the EHS [21]. We also investigated cross-sectional associations between the identified metabolite profile and domain-specific cognitive tests.

## Methods

### Study population

This is a cross-sectional analysis nested within the EHS, an ongoing population-based prospective cohort study conducted in the Epirus region in the Northwestern Greece. A detailed description of the study design and recruitment procedures have been published elsewhere [21]. From the initial sample of 1,596 participants recruited up to 15 October 2021, we excluded 60 participants with self-reported Alzheimer’s disease (n=1), Parkinson’s disease (n=2), epilepsy (n=11), Schizophrenia disease (n=1), major depression disorder (n=42) and bipolar disorder (n=3), 23 participants with missing data on cognitive scores (n=22), and MEDAS (n=1) and 263 without metabolomic measurements (**Figure 1**).

**Figure 1.**
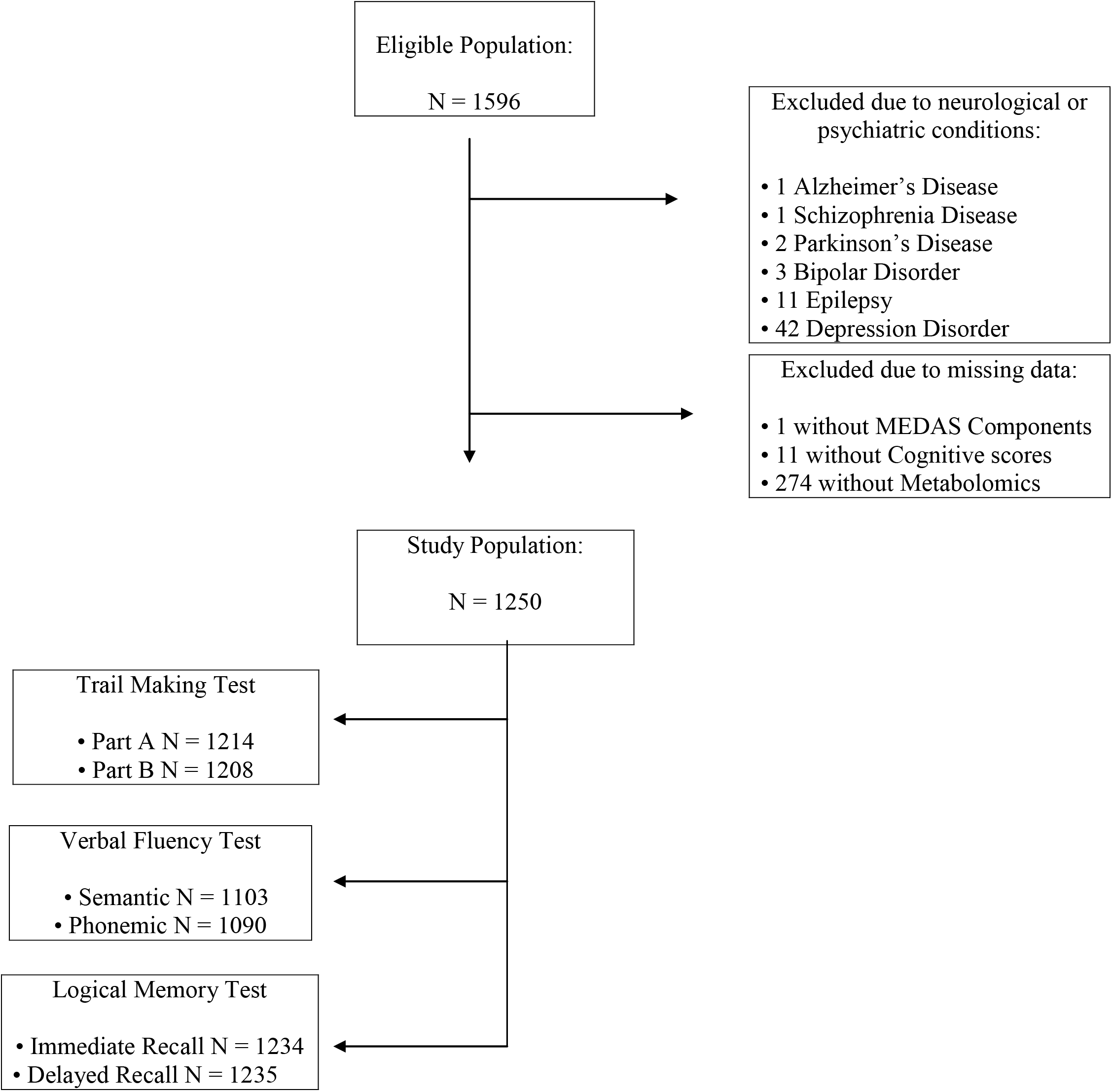
Flowchart of study participants. Abbreviations: MEDAS; Mediterranean Diet Adherence Screener.

The study was approved by the Research Ethics Committee of the University of Ioannina and was conducted in accordance with the ethical principles set forth by the Declaration of Helsinki [22]. All participants gave their written informed consent for participation in the study.

### Metabolomic profiling

Metabolomic analyses were conducted on consecutive fasting plasma ethylenediaminetetraacetic acid (EDTA) samples collected until October 2021 and stored at − 80°C. The samples were analyzed using the Nightingale Health’s NMR-based metabolomic profiling platform. This provides simultaneous quantification of 250 metabolites, including lipids, fatty acids, amino acids, ketone bodies, glycolysis-related metabolites, markers of inflammation and fluid balance as well as lipoprotein subclass distribution, particle size and composition. Details of the metabolite profiling platform and experimentation have been described previously [23-25]. The median and interquartile range of concentrations for the 250 quantified metabolites are given in **Supplementary Table S1**.

### Adherence to the Mediterranean diet

The degree of adherence to a traditional MedDiet was assessed using the 14-item Mediterranean Diet Adherence Screener (MEDAS) [26]. Fourteen questions on habitual intake of several food items are included in this screener including key components of the MedDiet such as olive oil, vegetables, fruits, nuts, red and white meat, butter, sweets, soft drinks and wine. Each of the 14 items is scored with 1 or 0, depending on whether participants adhere to each MedDiet component or not, respectively, and the resulting MEDAS score ranges from 0 (worst adherence) to 14 (best adherence).

### Cognitive performance measures

Trained personnel administered the Greek versions of the following neuropsychological tests: Trail Making Test, Verbal Fluency test and Logical Memory test.

The Trail Making Test (TMT) [27] is one of the most commonly used and well-established neuropsychological tests for detecting cognitive dysfunction, and consists of the TMT-A and TMT-B. In the TMT-A, which is related to cognitive domains such as visual scanning, attention, and processing speed, participants were asked to connect a set of dots with numbers that are randomly displayed on a piece of paper in an ascending order, as fast as possible. In the TMT-B, which associates with more complex cognitive abilities including executive function and processing speed, participants were asked to connect a set of dots with numbers and letters that are randomly distributed on a piece paper in an ascending order, with the added task of alternating between numbers and letters. Scoring was based on seconds needed to complete the test, with lower scores indicating better cognitive performances.

The Verbal Fluency test (VF) [27] is a measure of executive and verbal functioning. Participants were asked to generate as many words as possible, from either a category fluency (animals) or a letter fluency (starting with a specified letter), in 1 minute. The total score was the sum of the correct words.

The Logical Memory test (LM) [28] is a verbal episodic memory test. Participants were read a short story and asked to recall the story from memory immediately after (immediate recall). The procedure of immediate recall was repeated once, so that a learning curve was achieved. Approximately 20 minutes later, participants were asked again to recall the story (delayed recall condition). The number of correctly recalled information compiled the total score, ranging from 0–32 for the immediate recall condition and 0–16 for the delayed recall condition.

### Covariate Assessment

Information about demographic characteristics (i.e., age, sex, educational level) and lifestyle factors (i.e., recreational physical activity, smoking habits, alcohol consumption) was collected through a standard questionnaire, more details of which were previously described [21]. Body weight and standing height were measured using SECA equipment and body mass index (BMI) was calculated as weight divided by height squared (kg/m^2^).

### Statistical analyses

Characteristics of study participants are described as means and standard deviations (SD), or medians and interquartile ranges for quantitative variables, and percentages for categorical variables.

Two out of the 250 plasma metabolites, namely glycerol and β-hydroxybutyrate, were removed because of the high number of missing values (>20%) (**Supplementary Figure S1**). The remaining 248 metabolites had an average percentage of missingness (min, max) equal to 0.24% (0.24, 18.72%). Missing values of these metabolites were imputed using the random forest imputation approach (“missForest” function from the “missForest” R package), as recommended for metabolomics studies [29, 30]. The inverse normal transformation, which generates a rank-based standard normal distribution (mean=0, SD=1) was applied to the 248 metabolites [31].

To identify a metabolite profile for adherence to the MedDiet, we regressed MEDAS on the 248 NMR metabolites. Due to the high dimensionality and collinear nature of the data (**Supplementary Figure S2**), we performed Gaussian linear regression with elastic net penalty (implemented in the “glmnet” R package) [32], that combines the penalties from the least absolute shrinkage and selection operator (Lasso) and Ridge regressions. We evaluated the alpha parameter from 0.1 (we did not use Ridge regression [alpha=0] as not being able to perform variable selection) to 1 (Lasso regression) in ∼0.05 increments, together with the tuning parameter [λ (lambda)], using a 10-fold cross-validation (CV) approach, to identify the optimal combination regarding model accuracy and complexity. Specifically, we split 90% of the data selected randomly into training set and the remaining 10% sample into validation set. Then, we applied a 10-fold CV on the training set to find the best combination of alpha and lambda parameters keeping those with the lowest root mean-squared error (RMSE). Then, in the initial validation set we computed the RMSE and the Pearson correlation coefficient for these tuning parameters. Running this loop 10 times, in 7 out of 10 iterations, the alpha parameter had a value of 0.1 giving the lowest RMSE (**Supplementary Figure S3**). For this alpha value, we performed 200 times a 10-fold CV to our main data set to find 200 different values of lambda with the lowest RMSE, keeping the mean value. Finally, for lambda=0.0788 (95% confidence interval [CI]: 0.0777-0.0797) (**Supplementary Figure S4**) and with permutation test to find the 95% CI, we had the expected lambda with the lowest RMSE (**Supplementary Figure S5**). To obtain the metabolites coefficients, we randomly split the data 10 times to training and validation sets as before. From each of the 10 iterations, we applied these alpha and lambda values in each elastic net regression for every training set. Then, we built the metabolite model by keeping the mean value of those metabolites that were consistently selected on each iteration (i.e., metabolites selected 10 times). To estimate the 95% CI for each metabolite, we built a new metabolite model using 200 permutation tests. For the 200 different values of lambda, we calculated 200 different metabolite models. In each model, we kept only the mean value of the metabolites that was selected 10 times as before. Finally, we calculated the metabolite score as the weighted sum of the averaged coefficients from the 10 iterations for each selected metabolite. **Supplementary Figure S6** shows the penalized beta coefficients of metabolites in the model as functions of lambda.

Pearson correlation was calculated to evaluate the performance of the metabolite profile in assessing adherence to the MedDiet. To avoid overfitting, a 10-fold CV procedure was performed diving the complete dataset into training and validation sets (90% and 10%, respectively).

Associations of individual metabolites that with each food component and MEDAS were estimated using Spearman’s rank correlations. After merging the two olive oil and the two fruit components, a P-value = 0.05/(12*15) was considered as statistically significant for the metabolites-food components associations (Bonferroni correction for 12 MEDAS components x 15 independent clusters of metabolites) and P-value = 0.05/15 for the associations with MEDAS. The 15 independent clusters used in the correction, were identified using a data-driven unsupervised learning approach based on Ward’s minimum variance method that is applied in the hierarchical clustering.

The metabolite score was standardized by its z-score (mean=0; SD=1) before analyses. Multivariable linear regression models were fitted to investigate associations between the MEDAS score, the metabolite score associated with MEDAS and the continuous scores derived from the three neuropsychological cognitive tests and a composite z-score (cognitive battery) calculated by averaging the z-scores of every cognitive test. The models were adjusted for age (continuous), sex, education (primary and secondary school, high school, higher education), BMI (continuous), smoking status (current, former, never smokers), alcohol consumption (never, less than once/month, 1–3 times/month, 1–2 times/week, almost every day) and recreational physical activity (measured in Metabolic Equivalents of Energy Expenditure [METs] per hour/week; continuous). To test whether the metabolite score was associated with cognitive measures due to its correlation with MEDAS, we further adjusted for MEDAS z-score. To account for multiple testing, we adjusted P-values of the multivariable-adjusted associations with the use of the Benjamini-Hochberg false discovery rate (FDR) procedure [33]. A FDR-adjusted P-value < 0.05 was considered to be statistically significant after accounting for 7 tests corresponding to the 7 cognitive tests. All analyses were performed using R version 4.1.2 (2021-11-01) (R Foundation for Statistical Computing, Vienna, Austria).

## Results

Participants’ characteristics (59.6% female) are summarized in **Table 1**. The mean±SD age and BMI of the participants were 47.8±10.7 and 26.4±4.7, respectively. The majority of them had a high education level (63%). The mean±SD MEDAS was 7.3±1.7, suggesting a moderate adherence to the MedDiet.

**Table 1.** Characteristics of study participants.

| Characteristics | Total (N = 1250) |
| --- | --- |
| **Age (Years)** | 47.79 ± 10.71 |
| **BMI (Kg/m<sup>2</sup>)** | 26.72 ± 4.69 |
| **MEDAS (Score)** | 7.26 ± 1.70 |
| **Gender** |  |
| Female | 745 (59.6 %) |
| Male | 505 (40.4 %) |
| **Education** |  |
| Primary and Secondary School <sup>a</sup> | 95 (7.6 %) |
| High School <sup>b</sup> | 364 (29.12 %) |
| Higher Education <sup>c</sup> | 791 (63.28 %) |
| **Smoking Status** |  |
| Non-smokers | 562 (44.96 %) |
| Former smokers | 266 (21.28 %) |
| Current smokers | 422 (33.76 %) |
| **Alcohol Consumption** |  |
| Never | 160 (12.8 %) |
| Less than once/month | 383 (30.64 %) |
| 1-3 times/month | 202 (16.16 %) |
| 1-2 times/week | 356 (28.48 %) |
| 3-4 times/week | 91 (7.28 %) |
| Almost everyday | 58 (4.64 %) |
| **Physical activity (METs-hours/week)** | 9 [0,21] |
| **Cognitive performance scores** |  |
| Trail Making Test (Part A)<br>(seconds) | 33.87 [27.77,41.73] |
| Trail Making Test (Part B)<br>(seconds) | 61.55 [52.36,73.47] |
| Verbal Fluency (Semantic) (counts) | 22 [18,26] |
| Verbal Fluency (Phonetic) (counts) | 10 [7,13] |
| Logical Memory (Immediate Recall) (counts) | 23 [20,26] |
| Logical Memory (Delayed Recall) (counts) | 14 [12,15] |
| Cognitive Battery | 0.28 [-0.11,0.59] |

Mean ± standard deviation or median [interquartile range] and frequency (percentage) are presented for continuous and categorical variables, respectively.
Abbreviations: BMI, body mass index; MEDAS; Mediterranean Diet Adherence Screener; METs, Metabolic Equivalents of Energy Expenditure. <sup>a</sup> Elementary school or junior high school, up to 9 years of education. <sup>b</sup> High school, up to 12 years of education. <sup>c</sup> University degree/MSc/PhD/Postdoc, more than 13 years of education. There are 36 missing values for Trail Making Test (A), 42 for Trail Making Test (B), 147 for Verbal Fluency (Semantic), 160 for Verbal Fluency (Phonetic), 16 for Logical Memory (Immediate Recall), 15 for Logical Memory (Delayed Recall) and 9 for Cognitive Battery.

### Associations of metabolites with MEDAS

**Table 2** presents the 42 metabolites selected 10 times in the 10-CV of the elastic net regressions for MEDAS score, including 22 metabolites with positive coefficients and 20 with negative coefficients, which accounted for 12.1% of the total variance of MEDAS. Half of the selected metabolites for MEDAS were lipoproteins and lipoprotein subclass particles. Five high-density lipoprotein (HDL) particles, mainly large-sized and rich in esterified cholesterol, were positively, while two large-(rich in free cholesterol and phospholipids), one very large (rich in esterified cholesterol)- and one small-sized (rich in triglycerides) HDL particles were negatively associated with MEDAS score. Most very low-density lipoprotein (VLDL) particles, very small/small-sized but also very large/large-sized and rich in free cholesterol, triglycerides and phospholipids, were negatively associated with MEDAS score. Positive associations between extremely large/very large-sized VLDL particles rich in free cholesterol, phospholipids and triglycerides with MEDAS score were also observed. Two intermediate-density lipoprotein (IDL) particles rich in free cholesterol and phospholipids, and a medium-sized LDL particle rich in free cholesterol were associated with higher MEDAS score.

**Table 2.** Metabolites ranked from the highest to the lowest elastic net positive and negative regression coefficients for MEDAS score. Exposure contrast is per SD/z-score increase of the metabolite.

| Metabolites | $\beta$ (95% CI) | Metabolites | $\beta$ (95% CI) |
| --- | --- | --- | --- |
| **Omega-3 FA** | 0.178 (0.162, 0.202) | **Degree of unsaturation** | -0.304 (-0.335, -0.276) |
| **DHA** | 0.155 (0.139, 0.173) | **Glycoprotein acetyls** | -0.167 (-0.184, -0.151) |
| **TG/total lipids in large HDL** | 0.141 (0.112, 0.176) | **SFA/total FA** | -0.151 (-0.167, -0.136) |
| **MUFA** | 0.140 (0.128, 0.153) | **Isoleucine** | -0.141 (-0.158, -0.123) |
| **Acetoacetate** | 0.125 (0.111, 0.141) | **Free chol/total lipids in** | -0.122 (-0.150, -0.097) |

|  |  | **large HDL** |  |
| --- | --- | --- | --- |
| **IDL** | 0.109 (0.091, 0.127) | **Omega-6 FA** | -0.119 (-0.135, -0.101) |
| **Phospholipids in IDL** | 0.085 (0.070, 0.100) | **Omega-6/omega-3 FA** | -0.105 (-0.118, -0.094) |
| **Citrate** | 0.083 (0.071, 0.095) | **SFA** | -0.101 (-0.133, -0.077) |
| **Alanine** | 0.078 (0.067, 0.093) | **TG in small VLDL** | -0.099 (-0.133, -0.077) |
| **Free chol/total lipids in chylomicrons and extremely large VLDL** | 0.066 (0.046, 0.087) | **Phenylalanine** | -0.096 (-0.106, -0.085) |
| **Glucose** | 0.065 (0.054, 0.076) | **Cholesteryl esters/total lipids ratio in very large HDL** | -0.089 (-0.119, -0.064) |
| **Cholesteryl esters in HDL** | 0.063 (0.049, 0.076) | **Phospholipids/total lipids ratio in large HDL** | -0.083 (-0.098, -0.064) |
| **Cholesteryl esters in small HDL** | 0.062 (0.050, 0.076) | **Free cholesterol/total lipids ratio in very small VLDL** | -0.078 (-0.101, -0.060) |
| **Acetate** | 0.061 (0.049, 0.072) | **Free cholesterol/total lipids ratio in very large VLDL** | -0.077 (-0.097, -0.059) |
| **Glycine** | 0.059 (0.049, 0.071) | **Phospholipids/total lipids ratio in very small VLDL** | -0.068 (-0.085, -0.052) |
| **Phospholipids/total lipids ratio in very large VLDL** | 0.053 (0.033, 0.077) | **Total TG** | -0.061 (-0.080, -0.046) |
| **Cholesteryl esters/total lipids in large HDL** | 0.049 (0.033, 0.068) | **Free cholesterol/total lipids ratio in large VLDL** | -0.059 (-0.084, -0.041) |
| **Free chol in medium LDL** | 0.045 (0.028, 0.068) | **TG/total lipids ratio in small HDL** | -0.046 (-0.064, -0.033) |
| **TG/total lipids ratio in very large VLDL** | 0.044 (0.031, 0.058) | **PUFA/MUFA** | -0.036 (-0.048, -0.022) |
| **Lactate** | 0.034 (0.025, 0.043) | **Albumin** | -0.031 (-0.040, -0.021) |
| **Total lipids in HDL** | 0.032 (0.021, 0.042) |  |  |
| **Free chol/total lipids ratio in IDL** | 0.028 (0.019, 0.036) |  |  |

Abbreviations: Chol, cholesterol; DHA, docosahexaenoic acid; FA, fatty acids; HDL, high-density lipoprotein; IDL, intermediate-density lipoproteins; LDL, low-density lipoprotein; MEDAS, Mediterranean Diet Adherence Screener; MUFA, monounsaturated fatty acid; PUFA, polyunsaturated fatty acid; SFA, saturated fatty acid; TG, triglycerides, VLDL, very low-density lipoprotein; CI, Confidence Interval.

Among the total fatty acids (FA), positive associations with MEDAS score were observed for omega-3 FA, docosahexaenoic acid (DHA), monounsaturated fatty acids (MUFA) and the short-chain FA acetate, whereas negative associations were observed for total TG, saturated fatty acids (SFA), the ratio SFA to total FA, omega-6 FA, the ratio omega-6 to omega-3 FA, and the ratio polyunsaturated fatty acids (PUFA) to MUFA.

Several amino acids were also associated with MEDAS score including alanine (positive), glycine (positive), isoleucine (negative) and phenylalanine (negative). Albumin, the most abundant protein in plasma, was negatively associated with MEDAS score.

Metabolites involved in energy production such as acetoacetate, citrate, glucose and lactate were positively associated with MEDAS score

Higher glycoprotein acetyls, a marker of inflammation, were associated with lower score of MEDAS.

The Pearson correlation coefficient between MEDAS and the corresponding plasma metabolite profile was 0.35 (95% CI, 0.30 to 0.40, P-value = 5.5 × 10^−37^) (**Figure 2**), but there was variation in the metabolite profile among participants reporting the same MEDAS.

**Figure 2.**
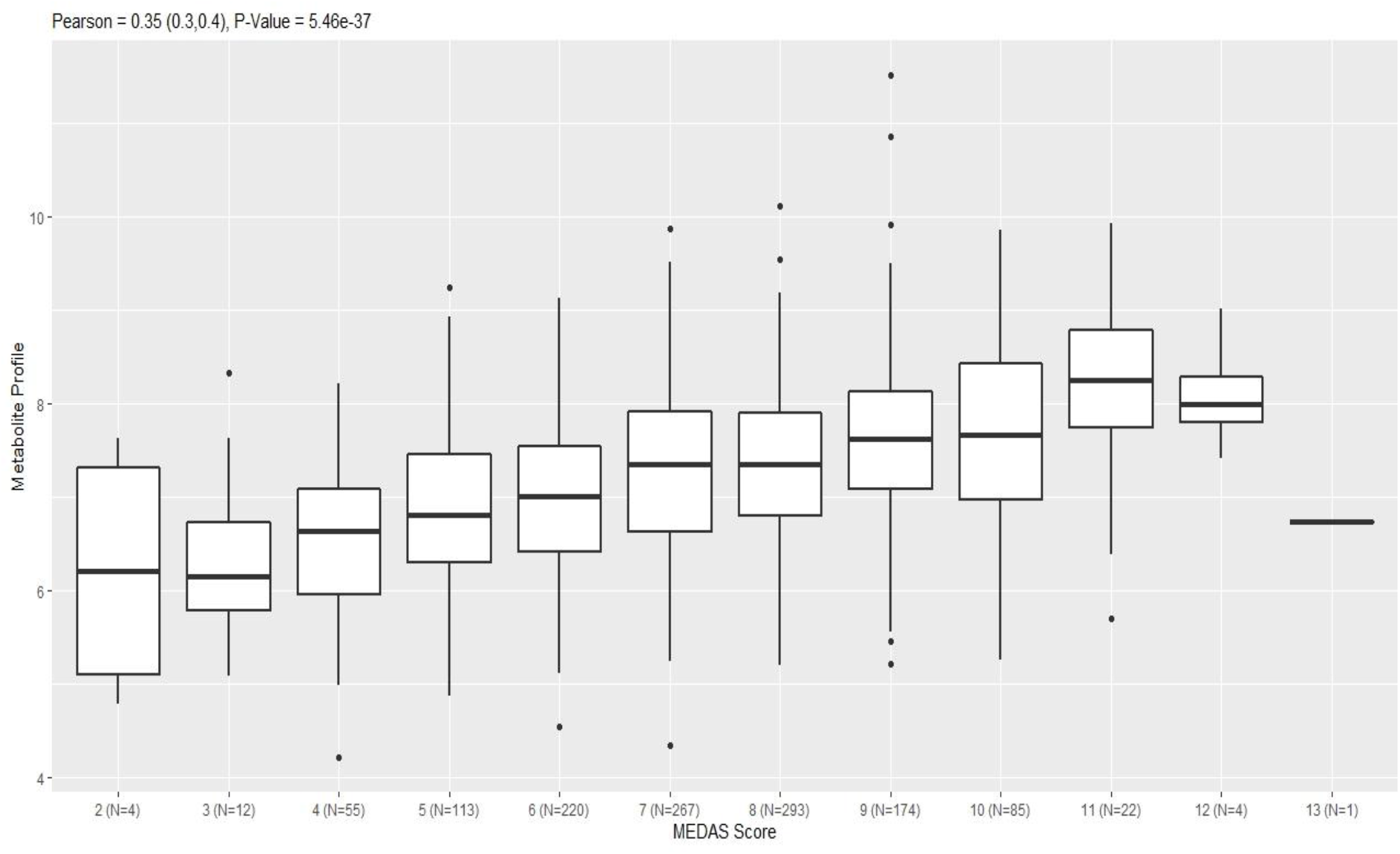
Pearson correlation between the Mediterranean Diet Adherence Screener (MEDAS) score and the metabolite profile.

Eighteen out of the 42 selected metabolites showed correlations with MEDAS score at P-value<0.05; Thirteen survived at a Bonferroni-corrected P-value (**Figure 3**). The associations were similar to those observed in **Table 2**. The MEDAS components correlated with the selected metabolites included poultry (m=18), sugar-sweetened beverages (m=16), fruits (m=14), vegetables (m=14), wine (m=15), olive oil (m=9), red meat (m=11), sweets (m=11), nuts (m=9), fish/seafood (m=8), butter/margarine (m=1), legumes (m=5) and sofrito (i.e. dishes with cooked vegetables and tomato sauce) (m=2) (**Figure 3**). However, the associations appeared to be stronger for intakes of fruits, red meat, sugar-sweetened beverages and sweets, and fish/seafood. For example, higher fish/seafood and fruits intake were positively correlated with omega-3 FA, while higher consumption of red meat was negatively correlated with cholesteryl esters and total lipids in HDL. Also, higher sugar-sweetened beverages intake was correlated with lower plasma concentrations of cholesteryl esters in HDL, and DHA.

**Figure 3.**
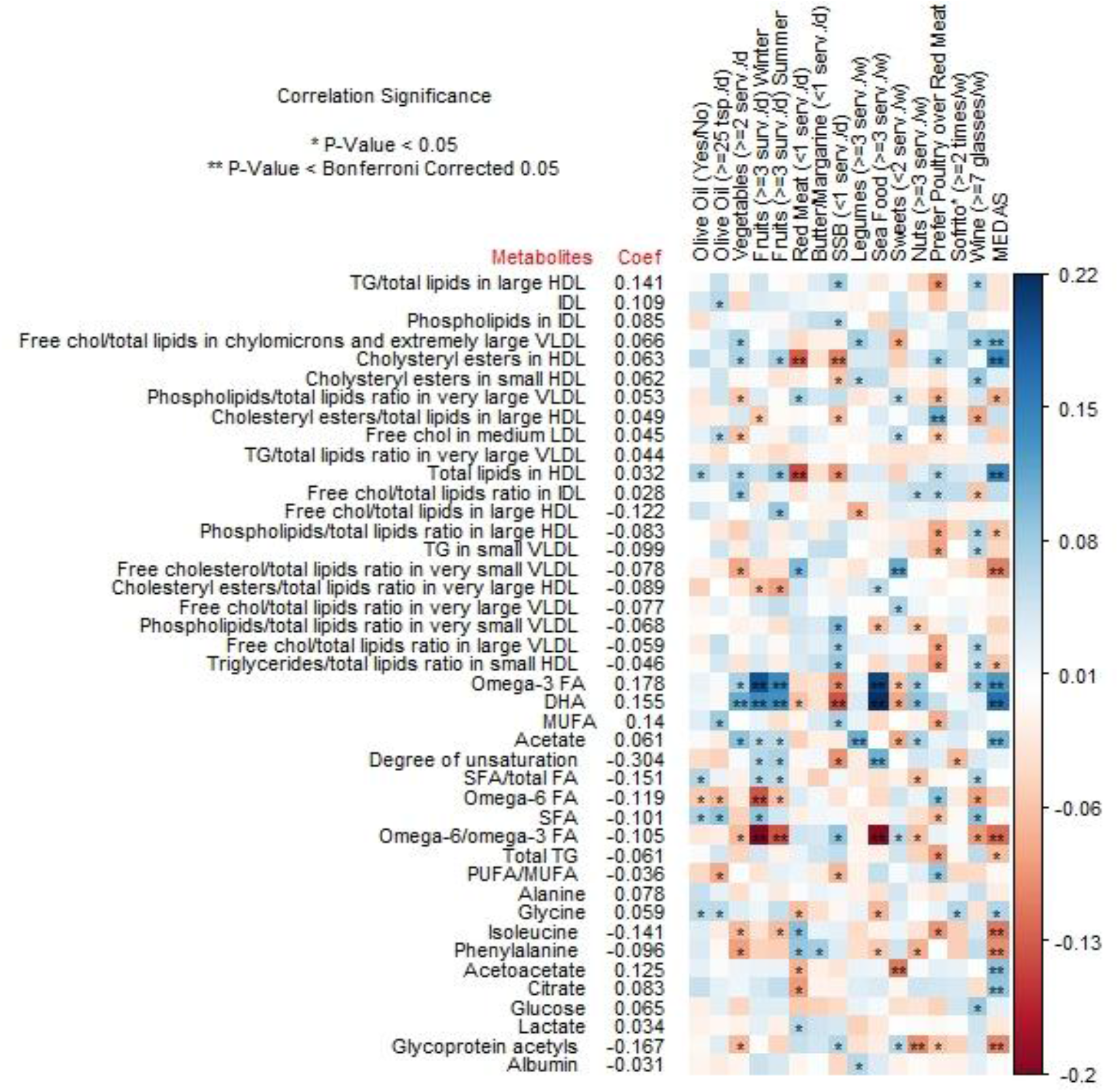
Spearman’s correlations of the 42 metabolites selected in the metabolite profile for MEDAS with MEDAS components and total MEDAS score. Presented from left to right are the metabolites’ coefficients obtained from elastic net regression (weights in the calculation of multi-metabolite score) and associations with MEDAS components and total MEDAS score. Abbreviations: Chol, cholesterol; DHA, docosahexaenoic acid; FA, fatty acids; HDL, high-density lipoprotein; IDL, intermediate-density lipoproteins; LDL, low-density lipoprotein; MEDAS, Mediterranean Diet Adherence Screener, MUFA, monounsaturated fatty acid; PUFA, polyunsaturated fatty acid; SFA, saturated fatty acid; TG, triglycerides, VLDL, very low-density lipoprotein; SSD, Sugar-Sweetened Beverages; *dishes with cooked vegetables and tomato sauce.

### Associations of MEDAS and metabolite score with cognitive tests

No significant associations between the scores of the metabolite profile and MEDAS with cognitive tests were observed (**Table 3, Supplementary Table S2**). Sensitivity analyses further adjusting for MEDAS z-score, self-reported supplement use, medication use, or history of CVD did not change the results (data not shown).

**Table 3.** Associations of the metabolite profile with cognitive performance scores. Exposure contrast is per SD/z-score increase of the metabolite score.

| Cognitive performance scores | Metabolic signature |  |  |  |  |  |
| --- | --- | --- | --- | --- | --- | --- |
|  | Model 1<sup>a</sup> |  |  | Model 2<sup>b</sup> |  |  |
| | $\beta$ | (95% CI) | P value<sup>1</sup> | $\beta$ | (95% CI) | P value<sup>1</sup> |
| **Trail Making Test (Part A) (seconds)** | -0.012 | (-0.69,0.666) | 0.972 | 0.054 | (-0.645,0.753) | 0.879 |
| **Trail Making Test (Part B) (seconds)** | 1.098 | (0.118,2.077) | 0.133 | 1.049 | (0.038,2.06) | 0.147 |
| **Verbal Fluency (Semantic) (counts)** | 0.191 | (-0.196,0.577) | 0.583 | 0.138 | (-0.262,0.539) | 0.697 |
| **Verbal Fluency (Phonetic) (counts)** | 0.089 | (-0.15,0.327) | 0.651 | 0.094 | (-0.154,0.342) | 0.697 |
| **Logical Memory (Immediate Recall) (counts)** | -0.162 | (-0.456,0.133) | 0.583 | -0.167 | (-0.471,0.137) | 0.656 |
| **Logical Memory (Delayed Recall) (counts)** | -0.162 | (-0.314,-0.009) | 0.133 | -0.169 | (-0.326,-0.011) | 0.147 |
| **Cognitive Battery** | -0.003 | (-0.038,0.032) | 0.972 | -0.004 | (-0.04,0.032) | 0.879 |

<sup>a</sup> Regressions were adjusted for age (continuous), sex, education (primary and secondary school, high school, higher education), body mass index (BMI; continuous), smoking status (current, former or never smokers), alcohol consumption (never, less than once/month, 1–3 times/month, 1–2 times/week, almost every day), recreational physical activity (measured in Metabolic Equivalents of Energy Expenditure (METs) per hour/week) (continuous).
<sup>b</sup> Regressions were adjusted for age (continuous), sex, education (primary and secondary school, high school, higher education), body mass index (BMI; continuous), smoking status (current, former or never smokers), alcohol consumption (never, less than once/month, 1–3 times/month, 1–2 times/week, almost every day), recreational physical activity (measured in Metabolic Equivalents of Energy Expenditure (METs) per hour/week) (continuous) and Mediterranean Diet Adherence Screener (MEDAS) score.
<sup>1</sup> Adjusted with the Benjamin-Hochberg False Discovery Rate method
Abbreviations: CI, Confidence Interval;

## Discussion

In the present study of almost 1250 Greek middle-aged adults, we identified 42 metabolites associated with MEDAS score. No associations between the identified metabolite profile and cognitive tests were observed.

The relationship between adherence to the MedDiet and blood metabolome has not been intensively explored. In the Mediterranean Diet in Northern Ireland trial, a metabolic profile of citric acid, pyruvic acid, betaine, mannose, acetic acid and myo-inositol showed modest discriminative accuracy for “low” *vs*. “high” MEDAS groups [18]. In the Prevention with Mediterranean Diet (PREDIMED) trial, participants following the MedDiet had a 1-year significant reduction in plasma lipid species [34], a finding that was also confirmed in two smaller MedDiet intervention studies [35, 36]. A recent cross-sectional analysis of MS metabolomic data from 1,859 participants within the PREDIMED trial identified a metabolic profile of adherence to the MedDiet and revealed that 67% of the identified plasma metabolites were lipid species involved in PUFA and lipid metabolic pathways [19]. To our knowledge, our study is the first to identify a metabolite profile of adherence to the MedDiet using an NMR-based metabolic profiling technique and regression with regularization. The identified metabolite profile composed of 42 metabolites, mainly lipoprotein subclasses and fatty acids. The profile was positively and significantly correlated with the self-reported measure of adherence to the MedDiet (r=0.35) and comparable to the degree of relationship between the metabolic profile and MEDAS score in the PREDIMED study (r=0.37). Similar to the PREDIMED study, we observed substantial variation in the metabolite profile among participants with the same MEDAS scores. Part of this inter-individual metabolic variability could be attributed to variations in genetics, microbiome and lifestyle [37, 38].

In our study, the MEDAS score was associated with metabolites related to most underlying MedDiet components replicating findings of previous studies and holding the promise that they could serve as candidate nutritional biomarkers. The positive correlations of omega-3 FA and especially DHA, and negative correlations of the ratio omega-6 to omega-3 with fish/seafood intake and MEDAS score were expected given the dietary origins of these FA and the high content of the MedDiet in PUFAs [19]. Also, acetate, a short-chain FA derived from fermentation of dietary fibers by gut microbes was correlated with higher intake of foods rich in fiber like vegetables, fruits, legumes and nuts and the Mediterranean dietary pattern [39].

The association between adherence to the MedDiet and lipoprotein particle subclass profile has been previously examined. A cross-sectional study conducted in 1,862 Irish adults revealed inverse associations between large VLDL, IDL, and small HDL and the Dietary Approaches to Stop Hypertension score but not with the MedDiet score [40]. In a small intervention study of 48 firefighters, participants following the MedDiet showed a decrease in large, medium and small LDL fractions such as total lipids, cholesterol, particle concentration or cholesterol esters [36]. In a larger intervention study of 296 participants from the PREDIMED trial, MedDiet increased the percentage of large HDL particles after 1-year [41]. In our study, higher adherence to the MedDiet could be related to decreased transfer of cholesteryl esters, possibly due to the decreased activity of cholesteryl ester transfer protein, and an increased ability of HDL to esterify cholesterol [41, 42]. Concerning the ratio of TG/total lipids in large HDL, it was reported that the increase in HDL particles in low HDL-C subjects is favoring the active ChoE/TG exchange process; TG enrichment of HDL notably increases the ability of hepatic lipase to remodel these HDL particles, resulting in release of lipid-poor apoA-I and enhanced clearance of HDL via the kidneys [43]. The distributional shift in cholesterol- and triglyceride-enriched VLDL particles with lower adherence to the MedDiet could be due to an elevation of plasma triglyceride levels and an effort to quickly repackage and remove them from the circulation [44]. Besides the correlations of fatty acids and lipoproteins with MEDAS components, the amino acids, isoleucine and phenylalanine were correlated with higher red meat consumption and a lower consumption of vegetables, fruits and the MEDAS score. Reductions in plasma branched-chain amino acids concentrations were observed after one year following the intervention with MedDiet+extra-virgin olive oil [45] and these amino acids were found inversely associated with healthy eating indices [17]. Consistent with a previous study [46], elevated glycine levels were negatively correlated with red meat consumption. Available evidence suggests elevated glycine utilization in response to a meat protein–based diet [47].

Acetoacetate, one of the main ketone bodies, was correlated with lower intakes of red meat and sweets, and with higher MEDAS score and we could speculate that a swift from glucose metabolism towards FA metabolism in those individuals following the MedDiet may have occurred [48]. On the other hand, in our study plasma lactate was positively correlated with red meat and butter; increased serum lactate was previously reported as a result of dairy products, as well as wheat bran, consumption [49]. The correlations of higher plasma citrate with higher fruit intake and adherence to the MedDiet confirms previous findings [18] suggesting a degree of specificity for this dietary pattern. Our finding on the negative correlations of glycoprotein acetyls with MEDAS score and healthy foods such as vegetables, and nuts, and positive correlations with sugary foods confirms previous results on the relationship between this low chronic inflammation marker and healthy eating [17]. Findings that increase in the MedDietScore may be associated with a decrease in serum albumin levels were also recorded previously [50].

Similar to our findings, a previous analysis from our research group using data from 1201 participants of the EHS cohort did not reveal associations between adherence to the MedDiet and cognitive performance [11]. Inconsistent results among studies examining this association exist. A lack of a beneficial effect of the MedDiet on cognitive ability was reported in observational and intervention studies [9, 10, 51], while others provided evidence of a potential protective role of the MedDiet on cognitive decline [5-8]. Clinical and methodological heterogeneity across studies could explain these inconsistencies. The discrepancies of the results among studies may be due to differences in geographical locations and characteristics (e.g. age, education) of the populations, application of different cognitive function tests and MedDiet adherence tools, or residual confounding due to lifestyle. Furthermore, the identified metabolite profile reflecting higher adherence to the MedDiet and capturing the metabolic effects of this dietary pattern in addition to other metabolic influences was not associated with cognitive performance.

Our study has several strengths. Our study participants were mostly middle-aged without comorbidities, which may affect the concentrations of metabolites. Furthermore, the use of regularized regression allowed us to conduct unbiased exploratory analysis and select biologically relevant metabolites, providing validity to our findings.

This study has some limitations that need to be mentioned. While studies addressing the metabolomic signature of MedDiet and cognitive function in middle-aged adults from the Mediterranean region are currently limited and our study may provide important insights for this association, the generalizability of our findings to older individuals from non-Mediterranean countries may be limited. Second, due to the cross-sectional design, causation and direction of causality cannot be inferred. More studies with repeated measures of metabolites and assessments of cognition are needed to understand how changes in MedDiet adherence relate to changes in metabolome, thereby influencing cognitive performance. Third, it is difficult to differentiate the metabolites that originate from the MedDiet and the metabolites that originate from other metabolic related factors. Finally, the use of a targeted metabolomic approach using an NMR platform may have partially covered the blood metabolome, limiting the identification of new metabolites associated with adherence to the MedDiet. Further research examining the associations of MedDiet adherence tools to a wider range of metabolites using untargeted metabolomics is needed.

In conclusion, our results extend previous research findings identifying a panel of 42 metabolites, measured by NMR, significantly correlated with adherence to the MedDiet. The identified metabolite profile showed no associations with cognitive performance in a healthy middle-aged Mediterranean population. Further studies are needed to evaluate external validity and examine associations between the MEDAS related metabolite profile and cognitive performance in other populations.

## Supporting information

Strobe

## Data Availability

Some of the data used in this manuscript may be available upon reasonable request to the authors given approval by the study's governing board.

## Author contributions

Conceptualization: CP1, EA, KKT

Data acquisition: MK, AK, GM

Statistical analyses: Christos Papagiannopoulos under the supervision of CP1 and KKT.

Findings interpretation: CP1 and KKT.

Writing – original draft: CP1

All the authors revised the manuscript for important intellectual content, and read and approved the final version.

