## Supplementary Files for "Mediterranean diet related metabolite profiles and cognitive performance in a Greek population"

**Supplementary Table S1**. Median and interquartile range of concentrations for the 250 metabolites quantified by nuclear magnetic resonance.

| Metabolites | ALL (N = 1250) |
|---|---|
| **Total-C (mmol/l)** | 5.074 [4.451,5.719] |
| **Non-HDL-C (mmol/l)** | 3.525 [2.959,4.147] |
| **Remnant-C (mmol/l)** | 1.571 [1.307,1.886] |
| **VLDL-C (mmol/l)** | 0.647 [0.498,0.816] |
| **Clinical-LDL-C (mmol/l)** | 2.841 [2.357,3.358] |
| **LDL-C (mmol/l)** | 1.943 [1.638,2.263] |
| **HDL-C (mmol/l)** | 1.491 [1.295,1.726] |
| **Total Triglycerides (mmol/l)** | 1.002 [0.778,1.383] |
| **VLDL-TG (mmol/l)** | 0.637 [0.458,0.966] |
| **LDL-TG (mmol/l)** | 0.140 [0.120,0.166] |
| **HDL-TG (mmol/l)** | 0.117 [0.095,0.146] |
| **Total-PL (mmol/l)** | 3.101 [2.804,3.396] |
| **VLDL-PL (mmol/l)** | 0.399 [0.299,0.529] |
| **LDL-PL (mmol/l)** | 0.663 [0.571,0.769] |
| **HDL-PL (mmol/l)** | 1.660 [1.479,1.888] |
| **Total-CE (mmol/l)** | 3.711 [3.281,4.173] |
| **VLDL-CE (mmol/l)** | 0.397 [0.311,0.496] |
| **LDL-CE (mmol/l)** | 1.414 [1.192,1.651] |
| **HDL-CE (mmol/l)** | 1.164 [1.012,1.349] |
| **Total-FC (mmol/l)** | 1.354 [1.175,1.541] |
| **VLDL-FC (mmol/l)** | 0.249 [0.186,0.326] |
| **LDL-FC (mmol/l)** | 0.527 [0.451,0.611] |
| **HDL-FC (mmol/l)** | 0.329 [0.283,0.382] |
| **Total-L (mmol/l)** | 9.254 [8.223,10.40] |
| **VLDL-L (mmol/l)** | 1.705 [1.279,2.323] |
| **LDL-L (mmol/l)** | 2.751 [2.349,3.193] |
| **HDL-L (mmol/l)** | 3.279 [2.885,3.742] |
| **Total-P (mmol/l)** | 0.018 [0.017,0.020] |
| **VLDL-P (mmol/l)** | <0.001 [<0.0001,0.0001] |
| **LDL-P (mmol/l)** | 0.001 [0.001,0.001] |
| **HDL-P (mmol/l)** | 0.016 [0.015,0.018] |
| **VLDL particle size (nm)** | 38.018 [37.236,38.937] |
| **LDL particle size (nm)** | 23.978 [23.906,24.030] |
| **HDL particle size (nm)** | 9.704 [9.569,9.868] |
| **Phosphoglycerides (mmol/l)** | 2.449 [2.202,2.691] |
| **TG/PG (ratio)** | 0.413 [0.323,0.554] |
| **Total cholines (mmol/l)** | 2.786 [2.523,3.036] |
| **Phosphatidylcolines (mmol/l)** | 2.276 [2.031,2.510] |
| **Sphingomyelins (mmol/l)** | 0.493 [0.448,0.537] |
| **ApoB (g/l)** | 0.881 [0.750,1.034] |
| **ApoA1 (g/l)** | 1.553 [1.413,1.719] |
| **ApoB/ApoA1 (ratio)** | 0.561 [0.463,0.678] |
| **Total fatty acids (mmol/l)** | 12.046 [10.711,13.697] |
| **Unsaturation (degree)** | 1.345 [1.312,1.376] |
| **Omega-3 (mmol/l)** | 0.396 [0.303,0.499] |
| **Omega-6 (mmol/l)** | 4.783 [4.325,5.298] |
| **PUFA (mmol/l)** | 5.201 [4.686,5.745] |
| **MUFA (mmol/l)** | 2.762 [2.385,3.288] |
| **SFA (mmol/l)** | 4.040 [3.559,4.611] |
| **LA (mmol/l)** | 3.793 [3.360,4.330] |
| **DHA (mmol/l)** | 0.206 [0.175,0.242] |

| | |
|---|---|
| **Omega-3 (%)** | 3.227 [2.678,3.892] |
| **Omega-6 (%)** | 40.124 [38.422,41.435] |
| **PUFA (%)** | 43.397 [41.876,44.557] |
| **MUFA (%)** | 23.046 [21.892,24.488] |
| **SFA (%)** | 33.577 [32.750,34.428] |
| **LA (%)** | 31.844 [30.145,33.427] |
| **DHA (%)** | 1.732 [1.482,1.982] |
| **PUFA/MUFA (ratio)** | 1.884 [1.717,2.030] |
| **Omega-6/Omega-3 (ratio)** | 12.335 [10.025,15.157] |
| **Alanine (mmol/l)** | 0.329 [0.287,0.379] |
| **Glutanine (mmol/l)** | 0.607 [0.559,0.661] |
| **Glycine (mmol/l)** | 0.237 [0.207,0.279] |
| **Histidine (mmol/l)** | 0.076 [0.069,0.083] |
| **Total BCAA (mmol/l)** | 0.375 [0.326,0.433] |
| **Isoleucine (mmol/l)** | 0.047 [0.038,0.057] |
| **Leucine (mmol/l)** | 0.109 [0.093,0.127] |
| **Valine (mmol/l)** | 0.219 [0.193,0.249] |
| **Phenylaline (mmol/l)** | 0.066 [0.059,0.074] |
| **Tyrosine (mmol/l)** | 0.059 [0.052,0.067] |
| **Glucose (mmol/l)** | 4.414 [4.048,4.782] |
| **Lactate (mmol/l)** | 2.308 [1.934,2.774] |
| **Pyruvate (mmol/l)** | 0.025 [0.016,0.038] |
| **Citrate (mmol/l)** | 0.059 [0.053,0.067] |
| **Glycerol (mmol/l)** | 0.094 [0.078,0.118] |
| **3-Hydroxybohbutyrate (mmol/l)** | 0.038 [0.022,0.076] |
| **Acetate (mmol/l)** | 0.020 [0.014,0.027] |
| **Acetoacetate (mmol/l)** | 0.020 [0.014,0.034] |
| **Acetone (mmol/l)** | 0.016 [0.014,0.020] |
| **Creatinine(µmol/l)** | 65.091 [57.596,74.614] |
| **Albumin (g/l)** | 42.486 [40.531,44.850] |
| **Glycoprotein acetyls (mmol/l)** | 0.774 [0.701,0.848] |
| **XXL-VLDL-P (mmol/l)** | <0.0001 [<0.0001,<0.0001] |
| **XXL-VLDL-L (mmol/l)** | 0.051 [0.014,0.157] |
| **XXL-VLDL-PL (mmol/l)** | 0.006 [0,0.023] |
| **XXL-VLDL-C (mmol/l)** | 0.021 [0.007,0.047] |
| **XXL-VLDL-CE (mmol/l)** | 0.014 [0.005,0.029] |
| **XXL-VLDL-FC (mmol/l)** | 0.007 [0.002,0.018] |
| **XXL-VLDL-TG (mmol/l)** | 0.025 [0.006,0.089] |
| **XL-VLDL-P (mmol/l)** | <0.0001 [<0.0001,<0.0001] |
| **XL-VLDL-L (mmol/l)** | 0.124 [0.068,0.213] |
| **XL-VLDL-PL (mmol/l)** | 0.022 [0.011,0.039] |
| **XL-VLDL-C (mmol/l)** | 0.039 [0.024,0.060] |
| **XL-VLDL-CE (mmol/l)** | 0.025 [0.016,0.036] |
| **XL-VLDL-FC (mmol/l)** | 0.014 [0.008,0.024] |
| **XL-VLDL-TG (mmol/l)** | 0.063 [0.032,0.114] |
| **L-VLDL-P (mmol/l)** | <0.0001 [<0.0001,<0.0001] |
| **L-VLDL-L (mmol/l)** | 0.245 [0.159,0.375] |
| **L-VLDL-PL (mmol/l)** | 0.046 [0.027,0.076] |
| **L-VLDL-C (mmol/l)** | 0.074 [0.047,0.111] |
| **L-VLDL-CE (mmol/l)** | 0.040 [0.026,0.059] |
| **L-VLDL-FC (mmol/l)** | 0.034 [0.021,0.052] |
| **L-VLDL-TG (mmol/l)** | 0.126 [0.084,0.194] |
| **M-VLDL-P (mmol/l)** | <0.0001 [<0.0001,<0.0001] |
| **M-VLDL-L (mmol/l)** | 0.541 [0.411,0.703] |
| **M-VLDL-PL (mmol/l)** | 0.125 [0.093,0.159] |
| **M-VLDL-C (mmol/l)** | 0.172 [0.131,0.221] |

| | |
|---|---|
| **M-VLDL-CE (mmol/l)** | 0.096 [0.072,0.123] |
| **M-VLDL-FC (mmol/l)** | 0.077 [0.057,0.097] |
| **M-VLDL-TG (mmol/l)** | 0.235 [0.177,0.324] |
| **S-VLDL-P (mmol/l)** | <0.0001 [<0.0001,<0.0001] |
| **S-VLDL-L (mmol/l)** | 0.370 [0.294,0.470] |
| **S-VLDL-PL (mmol/l)** | 0.093 [0.074,0.115] |
| **S-VLDL-C (mmol/l)** | 0.147 [0.115,0.184] |
| **S-VLDL-CE (mmol/l)** | 0.088 [0.069,0.113] |
| **S-VLDL-FC (mmol/l)** | 0.058 [0.046,0.072] |
| **S-VLDL-TG (mmol/l)** | 0.129 [0.099,0.172] |
| **XS-VLDL-P (mmol/l)** | <0.001 [<0.001,<0.001] |
| **XS-VLDL-L (mmol/l)** | 0.340 [0.287,0.403] |
| **XS-VLDL-PL (mmol/l)** | 0.099 [0.083,0.118] |
| **XS-VLDL-C (mmol/l)** | 0.179 [0.150,0.215] |
| **XS-VLDL-CE (mmol/l)** | 0.125 [0.105,0.150] |
| **XS-VLDL-FC (mmol/l)** | 0.054 [0.045,0.065] |
| **XS-VLDL-TG (mmol/l)** | 0.061 [0.050,0.075] |
| **IDL-P (mmol/l)** | <0.001 [<0.001,<0.001] |
| **IDL-L (mmol/l)** | 1.335 [1.141,1.532] |
| **IDL-PL (mmol/l)** | 0.316 [0.270,0.359] |
| **IDL-C (mmol/l)** | 0.923 [0.782,1.069] |
| **IDL-CE (mmol/l)** | 0.689 [0.582,0.797] |
| **IDL-FC (mmol/l)** | 0.235 [0.199,0.270] |
| **IDL-TG (mmol/l)** | 0.096 [0.082,0.113] |
| **L-LDL-P (mmol/l)** | 0.001 [0.001,0.001] |
| **L-LDL-L (mmol/l)** | 1.771 [1.513,2.046] |
| **L-LDL-PL (mmol/l)** | 0.390 [0.335,0.447] |
| **L-LDL-C (mmol/l)** | 1.286 [1.087,1.492] |
| **L-LDL-CE (mmol/l)** | 0.946 [0.801,1.097] |
| **L-LDL-FC (mmol/l)** | 0.337 [0.288,0.390] |
| **L-LDL-TG (mmol/l)** | 0.096 [0.082,0.112] |
| **M-LDL-P (mmol/l)** | <0.001 [<0.001,<0.001] |
| **M-LDL-L (mmol/l)** | 0.674 [0.561,0.797] |
| **M-LDL-PL (mmol/l)** | 0.178 [0.149,0.209] |
| **M-LDL-C (mmol/l)** | 0.465 [0.383,0.551] |
| **M-LDL-CE (mmol/l)** | 0.329 [0.268,0.394] |
| **M-LDL-FC (mmol/l)** | 0.134 [0.113,0.157] |
| **M-LDL-TG (mmol/l)** | 0.031 [0.026,0.037] |
| **S-LDL-P (mmol/l)** | <0.001 [<0.001,<0.001] |
| **S-LDL-L (mmol/l)** | 0.309 [0.265,0.354] |
| **S-LDL-PL (mmol/l)** | 0.097 [0.085,0.110] |
| **S-LDL-C (mmol/l)** | 0.197 [0.167,0.229] |
| **S-LDL-CE (mmol/l)** | 0.140 [0.119,0.164] |
| **S-LDL-FC (mmol/l)** | 0.057 [0.049,0.066] |
| **S-LDL-TG (mmol/l)** | 0.013 [0.011,0.016] |
| **XL-HDL-P (mmol/l)** | <0.001 [<0.001,<0.001] |
| **XL-HDL-L (mmol/l)** | 0.186 [0.143,0.250] |
| **XL-HDL-PL (mmol/l)** | 0.088 [0.062,0.123] |
| **XL-HDL-C (mmol/l)** | 0.093 [0.074,0.120] |
| **XL-HDL-CE (mmol/l)** | 0.068 [0.054,0.090] |
| **XL-HDL-FC (mmol/l)** | 0.025 [0.021,0.030] |
| **XL-HDL-TG (mmol/l)** | 0.006 [0.005,0.008] |
| **L-HDL-P (mmol/l)** | 0.002 [0.001,0.002] |
| **L-HDL-L (mmol/l)** | 0.773 [0.555,1.049] |
| **L-HDL-PL (mmol/l)** | 0.380 [0.277,0.510] |
| **L-HDL-C (mmol/l)** | 0.365 [0.256,0.509] |

| | |
|---|---|
| **L-HDL-CE (mmol/l)** | 0.284 [0.198,0.396] |
| **L-HDL-FC (mmol/l)** | 0.082 [0.058,0.114] |
| **L-HDL-TG (mmol/l)** | 0.026 [0.019,0.034] |
| **M-HDL-P (mmol/l)** | 0.004 [0.004,0.005] |
| **M-HDL-L (mmol/l)** | 1.117 [0.984,1.263] |
| **M-HDL-PL (mmol/l)** | 0.516 [0.459,0.580] |
| **M-HDL-C (mmol/l)** | 0.555 [0.481,0.639] |
| **M-HDL-CE (mmol/l)** | 0.457 [0.397,0.523] |
| **M-HDL-FC (mmol/l)** | 0.099 [0.084,0.115] |
| **M-HDL-TG (mmol/l)** | 0.043 [0.034,0.055] |
| **S-HDL-P (mmol/l)** | 0.010 [0.009,0.011] |
| **S-HDL-L (mmol/l)** | 1.190 [1.108,1.281] |
| **S-HDL-PL (mmol/l)** | 0.672 [0.626,0.725] |
| **S-HDL-C (mmol/l)** | 0.473 [0.439,0.508] |
| **S-HDL-CE (mmol/l)** | 0.350 [0.323,0.378] |
| **S-HDL-FC (mmol/l)** | 0.122 [0.114,0.132] |
| **S-HDL-TG (mmol/l)** | 0.043 [0.034,0.054] |
| **XXL-VLDL-PL (%)** | 13.274 [8.935,15.3] |
| **XXL-VLDL-C (%)** | 30.365 [24.622,41.597] |
| **XXL-VLDL-CE (%)** | 19.025 [14.686,27.758] |
| **XXL-VLDL-FC (%)** | 11.195 [9.520,13.625] |
| **XXL-VLDL-TG (%)** | 55.776 [45.988,62.768] |
| **XL-VLDL-PL (%)** | 17.495 [15.547,18.603] |
| **XL-VLDL-C (%)** | 30.741 [26.553,36.569] |
| **XL-VLDL-CE (%)** | 19.602 [15.571,24.523] |
| **XL-VLDL-FC (%)** | 11.297 [10.468,12.318] |
| **XL-VLDL-TG (%)** | 52.113 [46.332,56.723] |
| **L-VLDL-PL (%)** | 18.805 [16.574,20.037] |
| **L-VLDL-C (%)** | 28.746 [26.193,31.555] |
| **L-VLDL-CE (%)** | 15.587 [13.458,17.815] |
| **L-VLDL-FC (%)** | 13.297 [12.538,14.081] |
| **L-VLDL-TG (%)** | 52.969 [49.459,56.300] |
| **M-VLDL-PL (%)** | 22.60 [21.585,23.724] |
| **M-VLDL-C (%)** | 32.317 [28.221,35.810] |
| **M-VLDL-CE (%)** | 18.332 [15.153,21.008] |
| **M-VLDL-FC (%)** | 13.914 [12.924,14.919] |
| **M-VLDL-TG (%)** | 45.142 [40.398,50.252] |
| **S-VLDL-PL (%)** | 24.862 [23.728,26.131] |
| **S-VLDL-C (%)** | 39.501 [36.600,42.086] |
| **S-VLDL-CE (%)** | 23.805 [22.028,25.400] |
| **S-VLDL-FC (%)** | 15.506 [14.326,16.797] |
| **S-VLDL-TG (%)** | 35.592 [31.985,39.520] |
| **XS-VLDL-PL (%)** | 29.019 [28.407,29.530] |
| **XS-VLDL-C (%)** | 53.054 [50.720,55.272] |
| **XS-VLDL-CE (%)** | 37.126 [35.073,39.184] |
| **XS-VLDL-FC (%)** | 15.931 [15.631,16.183] |
| **XS-VLDL-TG (%)** | 17.960 [16.172,20.088] |
| **IDL-PL (%)** | 23.695 [23.141,24.142] |
| **IDL-C (%)** | 69.234 [67.867,70.305] |
| **IDL-CE (%)** | 51.597 [50.316,52.709] |
| **IDL-FC (%)** | 17.521 [16.958,18.029] |
| **IDL-TG (%)** | 7.198 [6.392,8.172] |
| **L-LDL-PL (%)** | 22.051 [21.708,22.411] |
| **L-LDL-C (%)** | 72.415 [71.521,73.155] |
| **L-LDL-CE (%)** | 53.368 [52.531,54.156] |
| **L-LDL-FC (%)** | 19.083 [18.54,19.608] |

| | |
|---|---|
| **L-LDL-TG (%)** | 5.436 [4.836,6.219] |
| **M-LDL-PL (%)** | 26.574 [26.088,27.012] |
| **M-LDL-C (%)** | 68.861 [67.968,69.514] |
| **M-LDL-CE (%)** | 48.547 [47.226,49.86] |
| **M-LDL-FC (%)** | 20.272 [19.27,21.117] |
| **M-LDL-TG (%)** | 4.640 [4.146,5.362] |
| **S-LDL-PL (%)** | 31.634 [30.766,32.473] |
| **S-LDL-C (%)** | 64.012 [62.971,64.781] |
| **S-LDL-CE (%)** | 45.278 [44.318,46.373] |
| **S-LDL-FC (%)** | 18.689 [17.847,19.269] |
| **S-LDL-TG (%)** | 4.338 [3.800,5.072] |
| **XL-HDL-PL (%)** | 46.963 [43.284,49.641] |
| **XL-HDL-C (%)** | 49.476 [47.468,52.38] |
| **XL-HDL-CE (%)** | 36.549 [35.414,38.157] |
| **XL-HDL-FC (%)** | 12.965 [11.703,14.776] |
| **XL-HDL-TG (%)** | 3.218 [2.475,4.553] |
| **L-HDL-PL (%)** | 48.89 [48.014,50.099] |
| **L-HDL-C (%)** | 47.826 [45.881,49.182] |
| **L-HDL-CE (%)** | 37.264 [35.541,38.436] |
| **L-HDL-FC (%)** | 10.608 [10.166,10.954] |
| **L-HDL-TG (%)** | 3.217 [2.490,4.369] |
| **M-HDL-PL (%)** | 46.203 [45.647,46.776] |
| **M-HDL-C (%)** | 49.909 [48.400,51.234] |
| **M-HDL-CE (%)** | 41.068 [39.754,42.236] |
| **M-HDL-FC (%)** | 8.848 [8.477,9.221] |
| **M-HDL-TG (%)** | 3.884 [3.025,4.902] |
| **S-HDL-PL (%)** | 56.611 [55.917,57.351] |
| **S-HDL-C (%)** | 39.809 [38.687,40.689] |
| **S-HDL-CE (%)** | 29.531 [28.461,30.414] |
| **S-HDL-FC (%)** | 10.239 [9.956,10.605] |
| **S-HDL-TG (%)** | 3.598 [2.958,4.397] |

Median [interquartile range].

Abbreviations: -C, cholesterol; -TG, triglycerides; -PL, Phospholipids; -CE, Cholesteryl esters; -FC, Free cholesterol; -L, Total lipids; -P, Lipoprotein particle concentrations; XXL-, Chylomicrons and extremely large; XL-, Very large; L-, Large; M-, Medium; S-, Small; XS- Very small; LA, Linoleic Acid; DHA, docosahexaenoic acid; FA, fatty acids; HDL, high-density lipoprotein; IDL, intermediate-density lipoproteins; LDL, low-density lipoprotein; MUFA, monounsaturated fatty acid; PUFA, polyunsaturated fatty acid; SFA, saturated fatty acid; VLDL, very low-density lipoprotein; VHDL, very high-density lipoprotein, %, Percentage.

**Supplementary Figure S1**. Plot of the 250 metabolites according to the percentage of missingness in 1250 participants.

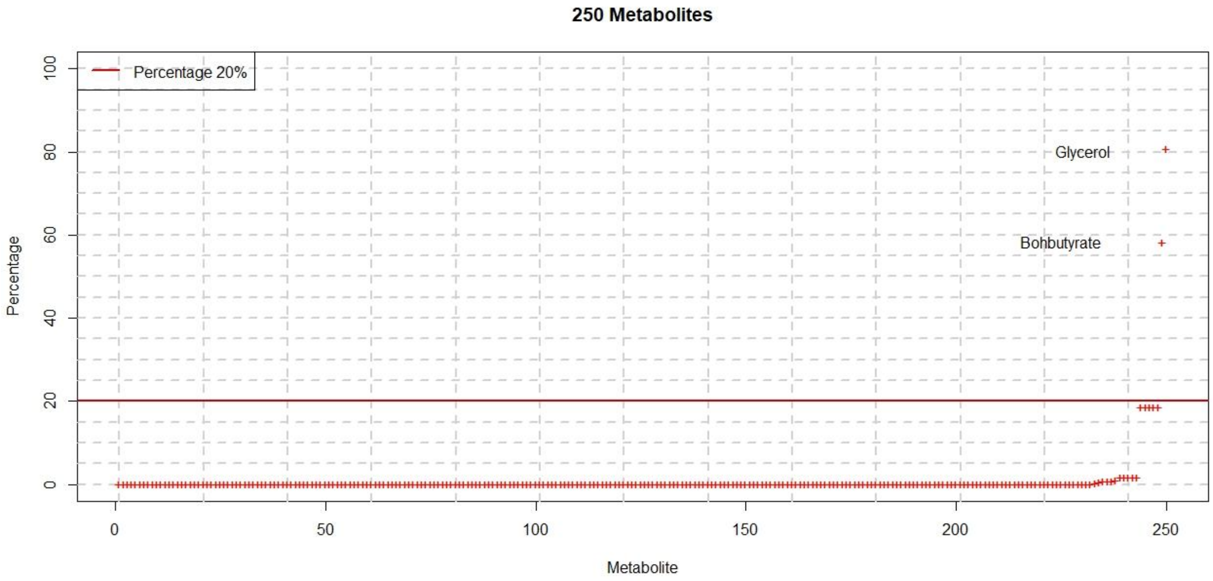


**Supplementary Figure S2**. Correlation matrix for all the 248 metabolites considered in the analysis. Colors represent directions of the correlation (blue-positive and red-inverse) while the color depth indicates correlation magnitudes (the darker the stronger).

**Supplementary Figure S3**. Plot of the lambda and Root Mean Square Error for different alpha values after the 10 iterations with the elastic net penalty. The colored dots are the alpha values.

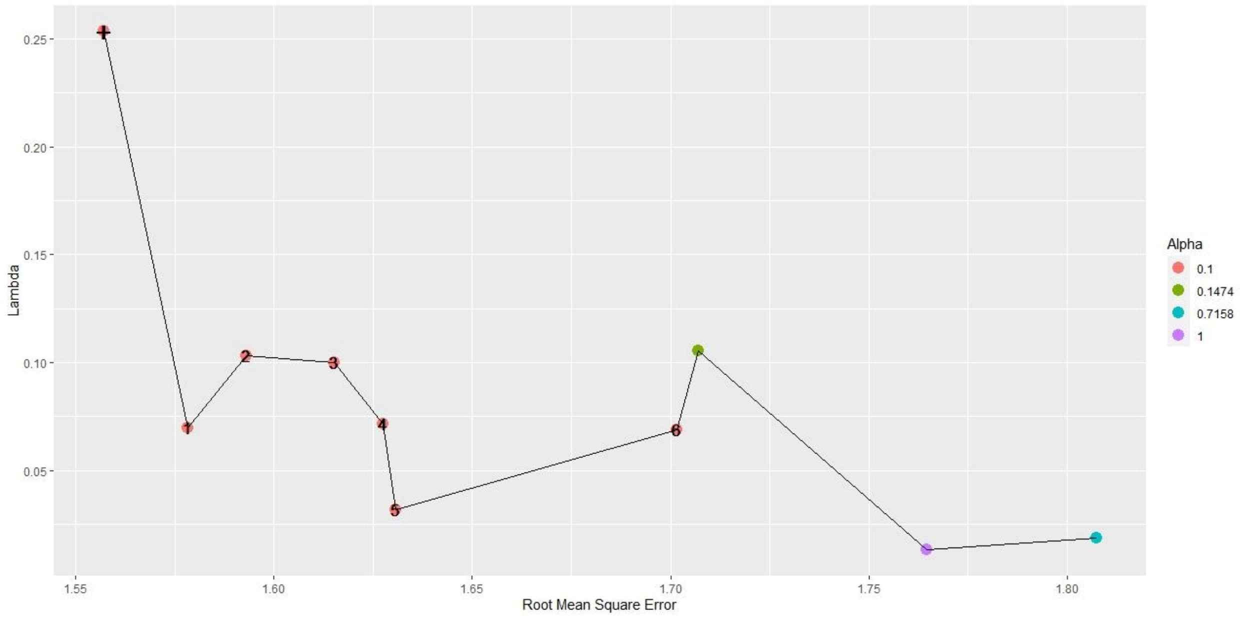

**Supplementary Figure S4**. Plot of the 200 simulated identified lambda for Alpha = 0.1. As the simulations increase the mean lambda converges to its value 0.0788.

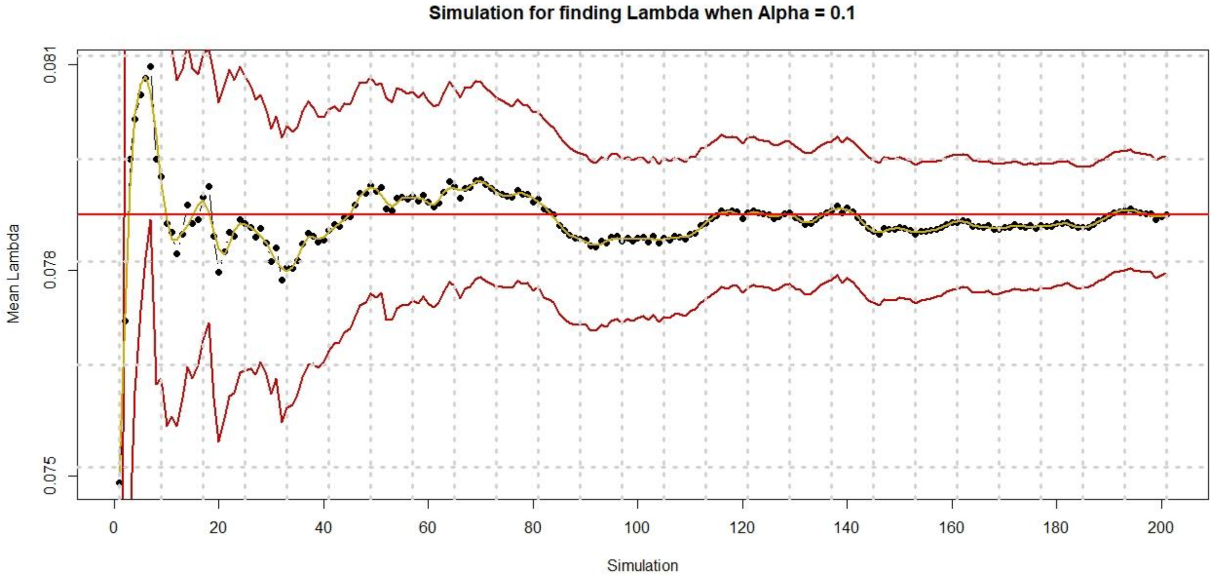


**Supplementary Figure S5**. Plot of the Mean-Squared Error as functions of log(λ) for the 200 times 10-fold cross-validation regression analyses with the elastic net penalty. The orange line is the mean form the cross-validation and the shaded area represent 95% range confidence interval.

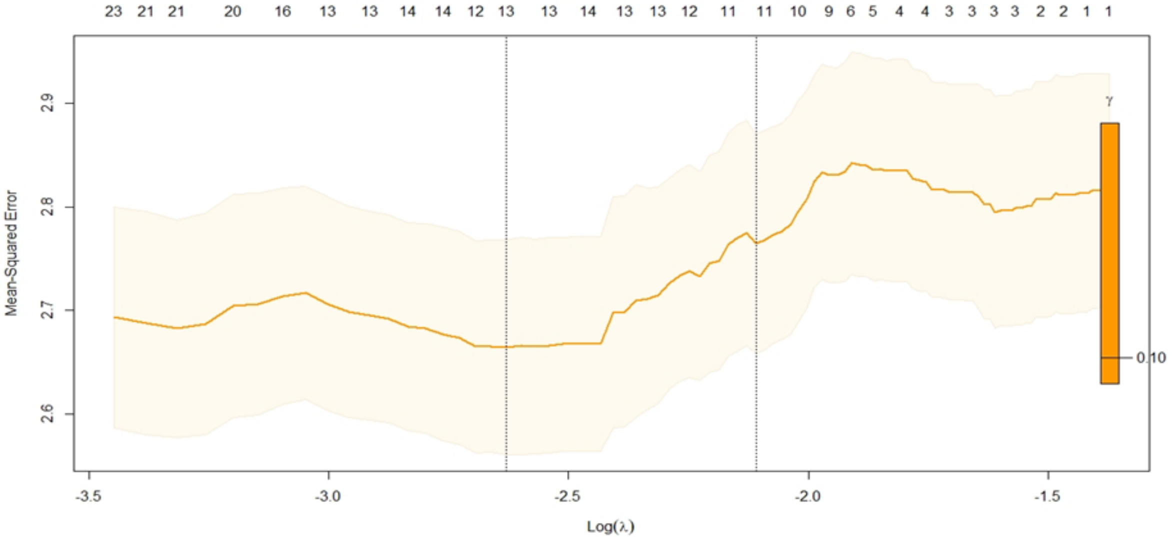

**Supplementary Figure S6**. Plot of the beta coefficients of metabolites in the model as functions of Lambda for the 10-fold cross-validation analyses with the elastic net penalty.

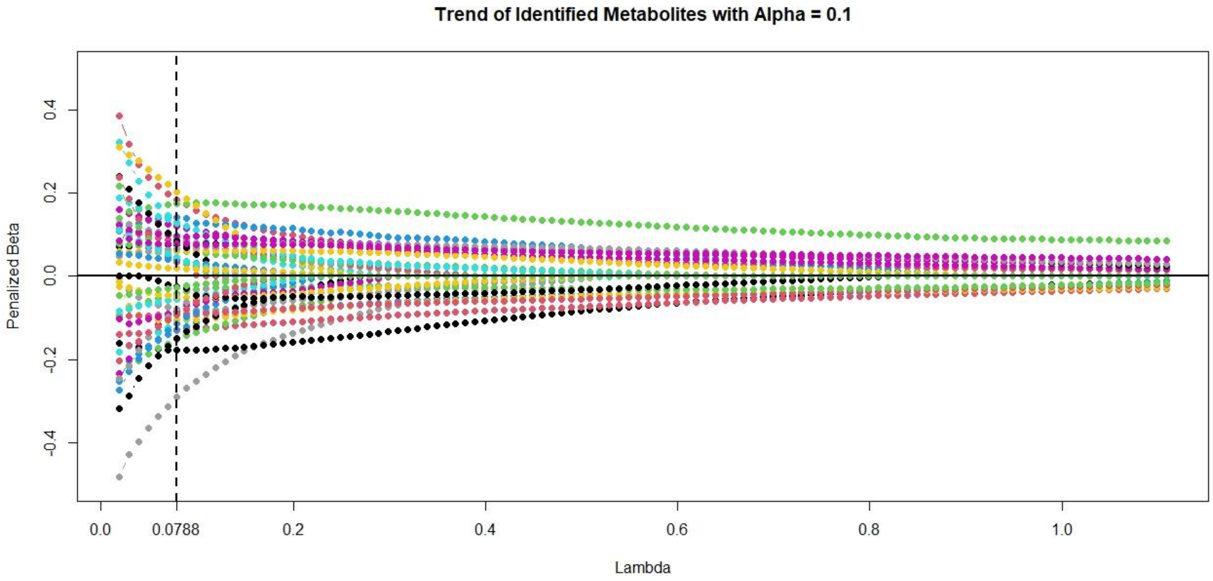

**Supplementary Table S2**. Associations of MEDAS with cognitive performance scores.

| Cognitive performance scores. | MEDAS score | | |
|---|---|---|---|
| | Model | | |
| | β | (95% CI) | P-Value |
| **Trail Making Test (Part A) (seconds)** | -0.144 | (-0.525,0.237) | 0.459 |
| **Trail Making Test (Part B) (seconds)** | 0.257 | (-0.297,0.81) | 0.363 |
| **Verbal Fluency (Sematic) (counts)** | 0.133 | (-0.085,0.351) | 0.232 |
| **Verbal Fluency (Phonetic) (counts)** | 0.004 | (-0.131,0.139) | 0.954 |
| **Logical Memory (Immediate Recall) (counts)** | -0.011 | (-0.177,0.155) | 0.895 |
| **Logical Memory (Delayed Recall) (counts)** | -0.007 | (-0.093,0.079) | 0.875 |
| **Cognitive Battery** | 0.001 | (-0.018,0.021) | 0.888 |

Regressions were adjusted for age (continuous), sex, education (primary and secondary school, high school, higher education), body mass index (BMI; continuous), smoking status (current, former or never smokers), alcohol consumption (never, less than once/month, 1–3 times/month, almost every day), recreational physical activity (measured in Metabolic Equivalents of Energy Expenditure (METs) per hour/week) (continuous).

Abbreviations: CI, Confidence Interval; MEDAS, Mediterranean Diet Adherence Screener.
