## Supplementary figures and images for "Mediterranean diet related metabolite profiles and cognitive performance in a Greek population"

### Supplementary Figure S2

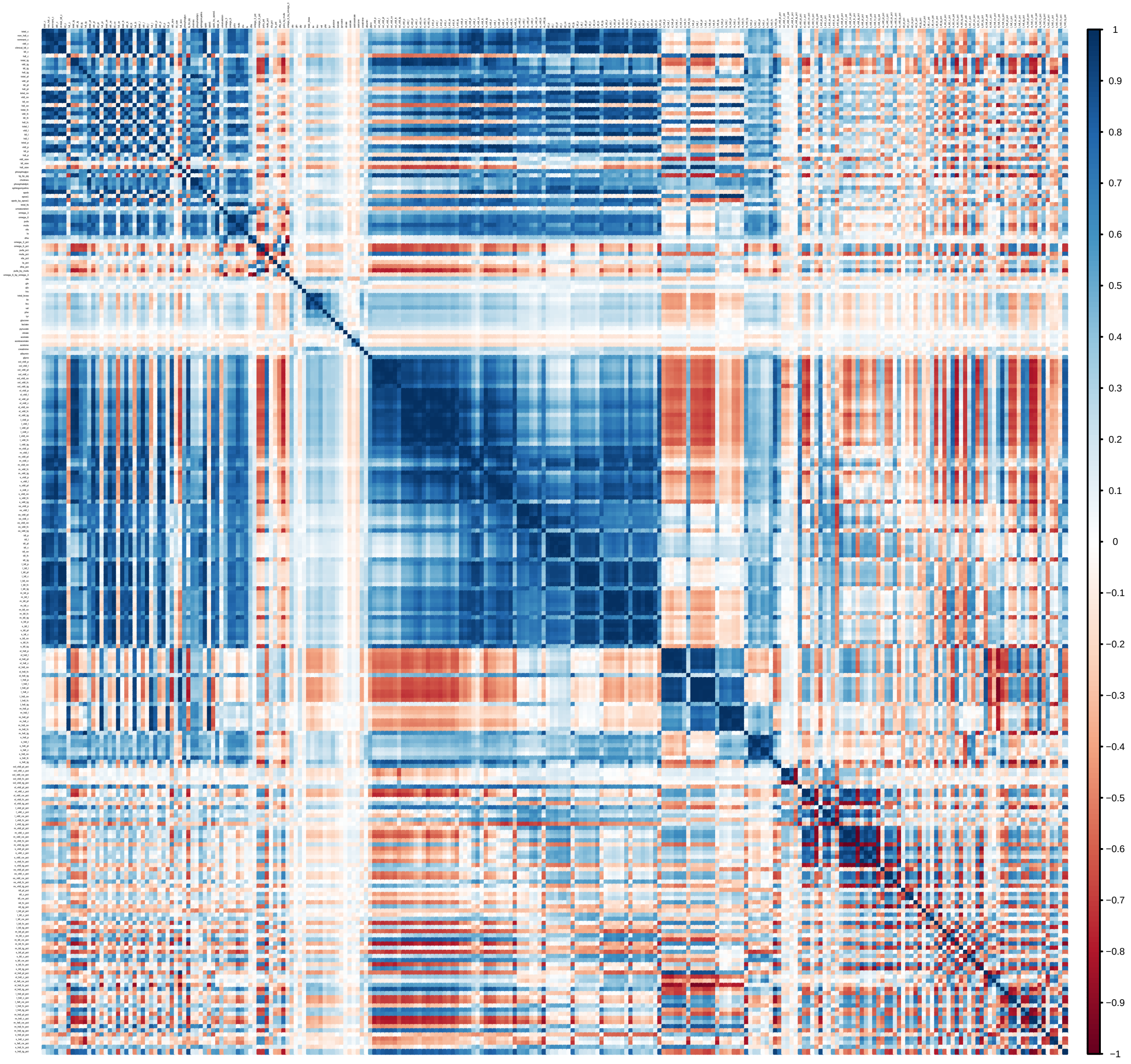
